# Predicting the COVID-19 epidemic in Algeria using the SIR model

**DOI:** 10.1101/2020.04.25.20079467

**Authors:** Mohamed Samir Boudrioua, Abderrahmane Boudrioua

## Abstract

The aim of this study is to predict the daily infected cases with Coronavirus (COVID-19) in Algeria. We apply the SIR model on data from 25 February 2020 to 24 April 2020 for the prediction. Following Huang et al (12), we develop two SIR models, an optimal model and a model in a worst-case scenario COVID-19. We estimate the parameters of our models by minimizing the negative log likelihood function using the Nelder-Mead method. Based on the simulation of the two models, the epidemic peak of COVID-19 is predicted to attain 24 July 2020 in a worst-case scenario, and the COVID-19 disease is expected to disappear in the period between September 2020 and November 2020 at the latest. We suggest that Algerian authorities need to implement a strict containment strategy over a long period to successfully decrease the epidemic size, as soon as possible.

## 1 Introduction

On 31 December 2019, a cluster of cases of pneumonia of unknown cause was detected in Wuhan City, Hubei Province of China (26). The Chinese authorities identify on 7 January the coronavirus disease (COVID-2019) as the causative virus (26). This epidemic continues to cause infections in other Chinese cities and in multiple countries around the world (24). On 01 April 2020, the World Health Organization (WHO) has reported a total of 823611 confirmed cases of COVID-19 infection with 40534 death cases worldwide (27). Algeria has reported the first case of COVID-19 on 25 February 2020, in Ouargla region and he was an Italian national (8, 1). Two other cases were reported on 01 march 2020 in Blida region in the North of Algeria (8, 1). The epidemic continues to spread in other regions of the country. On 03 April 2020, the Algerian authorities have declared 1171 confirmed cases with 105 deaths cases (1).

Various earlier studies have predicted the confirmed cases of COVID-19 infection in different countries. Kuniya (17) applied the well-known SEIR compartmental model to predict the epidemic peak of COVID-19 disease in Japan. He used data of daily reported cases of COVID-19, from15 January to 29 February 2020. He founds that the epidemic peak in Japan could possibly reach the early-middle summer. Roosa et al (25) forecast the covid-19 epidemic in China over a short-term period using generalized logistic growth model, the Richards growth model, and a sub-epidemic wave model. They employed data of cumulative confirmed cases between January 22, 2020 and February 9, 2020. They found that the epidemic has reached saturation in Hubei and other provinces of China. Hamidouche (8), introduce the Alg-COVID-19 Model to predict cumulative cases of COVID-19 in Algeria. This model allows predicting the incidence and the reproduction number R0 in the upcoming months (8). According to his results, the number of infected cases in Algeria will exceed 5000 on 07 April 2020 and it will double to 10000 on 11 April 2020 (8).

This study applies the Susceptible-Infected-Removed (SIR) model without demographics (no births, deaths, or migration) (15, 5, 14) to predict the daily confirmed cases of COVID-19 infection in Algeria. We use data of reported confirmed cases between February 25, 2020 and April 24, 2020. We predict the epidemic peak and the end-date of this disease using an optimal SIR model and an SIR model in a worst-case COVID-19 scenario. These results could be helpful for decision makers in Algeria to implement their future counter strategies to COVID-19 outbreak.

## 2 Method

### 2.1 Model

The SIR model without demographics is given by the following non linear system of ordinary differential equations (15, 14, 9):

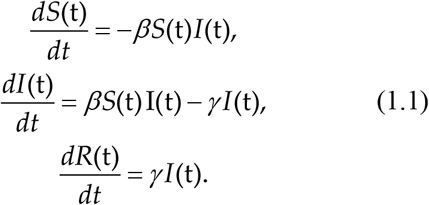

The function *S*(t) represents the number of susceptible cases at time t. The function *I*(t) represents the number of infected cases at time t, and the function *R*(t) is the compartment of removed cases from the disease, where:

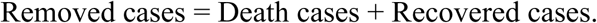

The parameters *β* and γ are called the contact rate and the removed rate, respectively. The initial condition of this model *S*(0), *I*(0), *R*(0) must satisfy the condition *S*(0) + *I*(0) + *R*(0) = *N*, where N is the population size (9).

The model can be written also as follows:

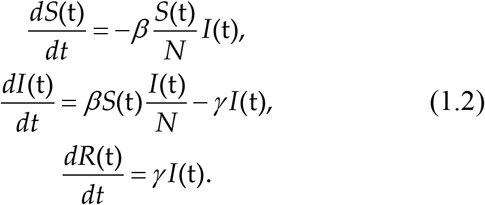

With: 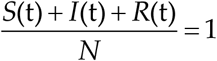 and S(0) > 0,I(0) > 0, *R*(0) = 0. (14)

The basic reproductive ratio *R*_0_ is’’ the average number of secondary cases arising from an average primary case in an entirely susceptible population, and it’s given by β/γ’’ (14). We integrate numerically our non-linear system using the Livermore Solver for Ordinary Differential Equations (LSODA) algorithm (10, 21). This algorithm handles stiff and non stiff systems of size N, with general form *dy* / *dt* = *f* (*t, y*). It has an automatic method selection where it uses Adams methods with the non stiff problems and Backward Differentiation Formula (BDF) methods with the stiff problems (10).

Following Huang et al (12), we consider two SIR models: The first one is an optimal model, where we suppose that the initial population size N is equal to the total of confirmed cases in Algeria.

In the second SIR model we assume that whole real population in Algeria is susceptible, with high contact rate *β* and high removed rate γ (either recovered or died). This is the worst-case scenario, and this model could be used to predict the largest infections of the epidemic (1212). Python software version 3.6.4 was used to implement the SIR model and solve it.

### 2.2 Maximum likelihood estimation

After resolving the SIR system we obtain the approximate functions 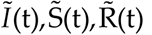. We are interested in the function *Ĩ*(t) the approximate of the function of the number of infections *I*(t) Since we have count data *y*_*t*_ (the real numbers of daily infected cases of COVID-19 in Algeria), we suppose that *y* follows a Poisson distribution with parameter 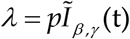 (16) :

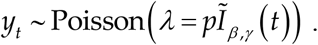

Where the parameter *p* is a constant reflects a combination of sampling efficiency and the detectability of infections (16). We ignore the constant *p in* this study.

Thus the likelihood function is defined as follows:

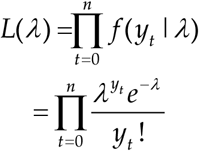

Then the log likelihood function is:

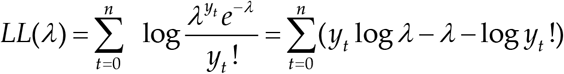

Ignoring the constant involving log *y*_*t*_ !, and substituting 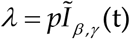, we get:

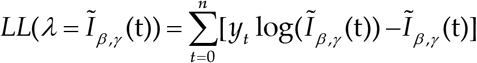

The objective is estimating the parameters *β* and γ that give us the optimum solution *Ĩ*(t). The Nelder-Mead algorithm (18) was used to find the minimum of the negative log likelihood function, and therefore maximize the log likelihood function. This algorithm is used for solving the unconstrained optimization problem based on the iterative update of a simplex:

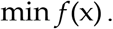

Where *f*: ℝ ^*n*^ → ℝ is the objective function and *n* the dimension (7, 2).

For more details about the Nelder-Mead algorithm, see (7, 2, 18).

The procedure of estimation can be resumed in the following two steps (19):

Step 1: Select the initial parameters *β*, γ and initialize the SIR model and the Nelder-Mead method,

Step 2: Run the Nelder-Mead algorithm to get new parameters which will be used in the simulation and the prediction of the COVID-19 epidemic.

All the calculations were conducted as previously stated by Python software version 3.6.4, following (6). The implementation codes can be found at https://github.com/MSamir9/Predicting-the-COVID-19-epidemic-in-Algeria-using-the-SIR-model

## 3 Empirical results

The data were obtained from Johns Hopkins University, Center for systems science and engineering (11). We used the number of daily new confirmed cases for the COVID-19 epidemic in Algeria, from 25 February 2020 to 24 April 2020.

Firstly we fit the optimal SIR model. We used equation (1.2) with initial parameters *β* = 0.3, *γ* = 0.1 and initial conditions:

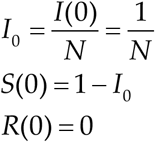

With *N* = 3127, equal to the total confirmed cases till 24 April 2020. Solving the SIR system and following the procedure of estimation cited previously, we get the new parameters: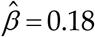 and 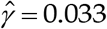. Thus the basic reproduction number *R*_0_ is given by:

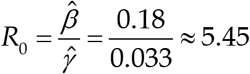

We substitute the new parameters in the model to simulate and predict the COVID-19 epidemic in Algeria.

**Figure 1** shows the predictions of the number of daily new infected cases with COVID-19 in Algeria, by fitting the optimal SIR model. The mean absolute error (MAE) is used to evaluate the accuracy of this model, where:

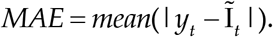

*y*_*t*_ is the real reported value at time t and *Ĩ*(t) is the predicted value at the same time t (13, 3). We get *MAE* = 14 as accuracy of the optimal SIR model for the prediction of new infected cases. Concerning the second model in a worst-case scenario COVID-19, we reset the initial parameters as follow:

**Figure 1:**
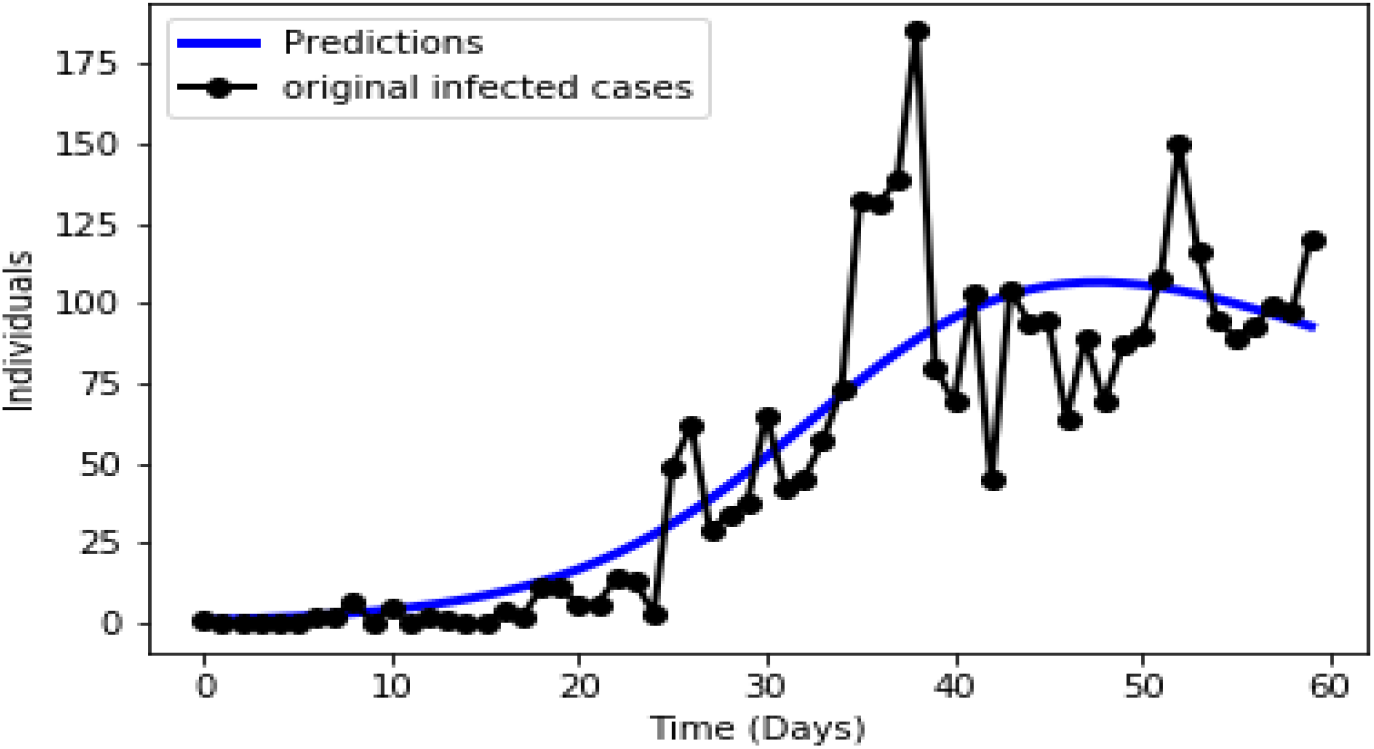
Fit of the optimal SIR model for Algeria infected cases with covid-19, from 25 February 2020 to 24 April 2020.

*β* = 0.95, as a high contact rate.

*γ* = 0.75, as a high removed rate (either recovered or death cases).

*N* = 43851044 The real population size of Algeria (28).

We proceed as before, we get the estimated parameters 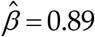, 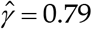, thus *R*_0_ ≈ 1.13.

**Figure 2** shows the simulation of infected cases with COVID-19 in a worst-case scenario. We get *MAE* = 30 as accuracy of this model. We remark that the predicted number of infected cases with COVID-19 in Algeria increase exponentially fast in this case.

**Figure 2:**
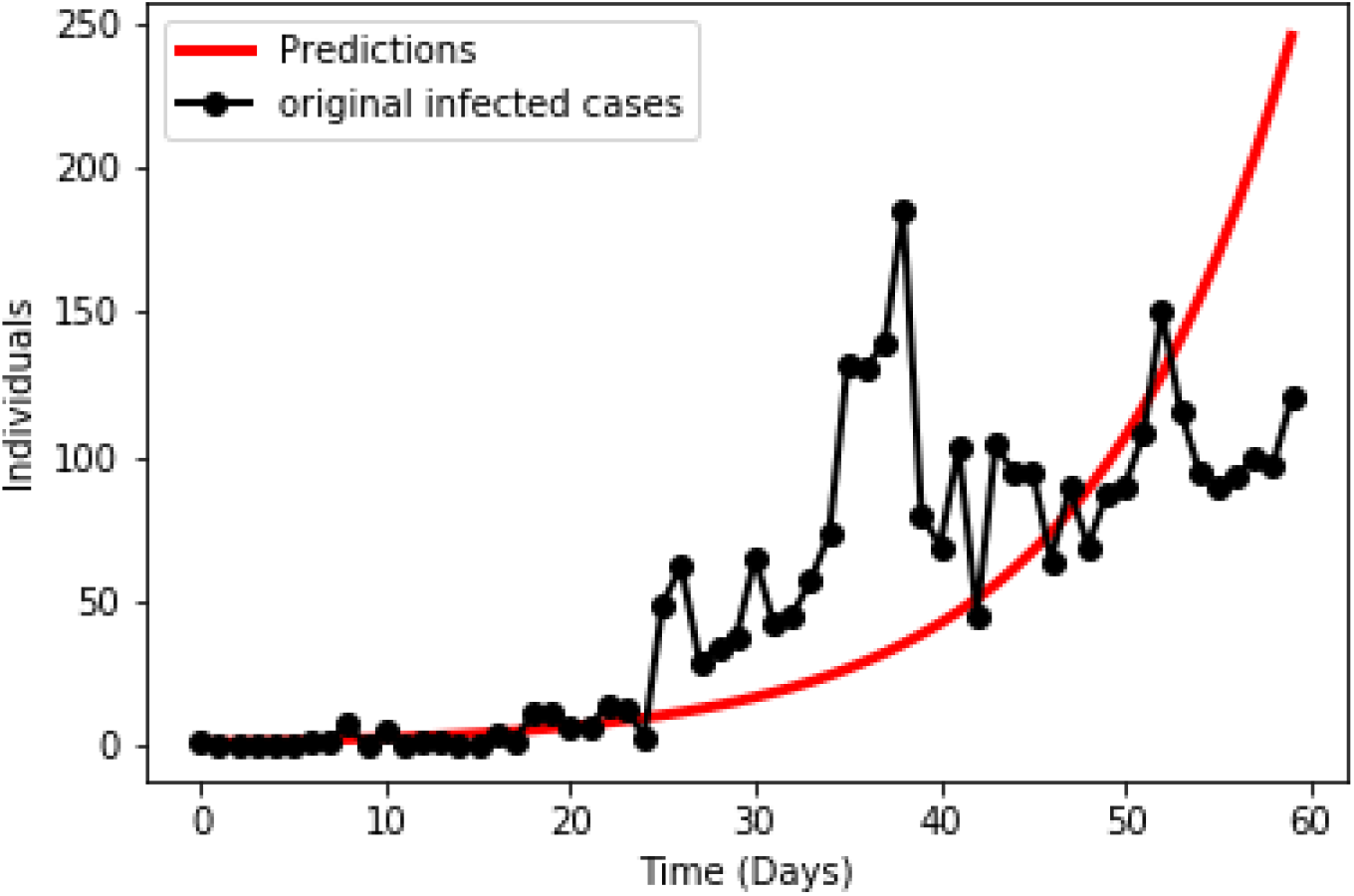
Fit of SIR model for Algeria infected cases in a worst-case scenario covid-19, from 25 February 2020 to 24 April 2020.

The prediction of the epidemic peak of COVID-19 in Algeria using both models is shown in **Figure 3**. We extract from this Figure that the predicted epidemic peak would be around 25 July 2020 at the latest, with about 2444 ×10^2^ infections, in a worst-case scenario. The predicted epidemic peak based on the optimal SIR model was on 13 April 2020 with 106 infections. The end-date of COVID-19 in Algeria is estimated to be around 16 November 2020 in a worst-case scenario, and around 18 September 2020 based on the optimal SIR model predictions. We can remark easily the large differences in the predicted infections between the two models. These differences represent the potentially preventable infections as a result of containment strategies (12). This shows the importance of containment strategies and how many COVID-19 infections can be prevented by these strategies. Stand on these results; we conclude that the COVID-19 epidemic will continue to spread in Algeria for about six upcoming months, at the latest.

**Figure 3:**
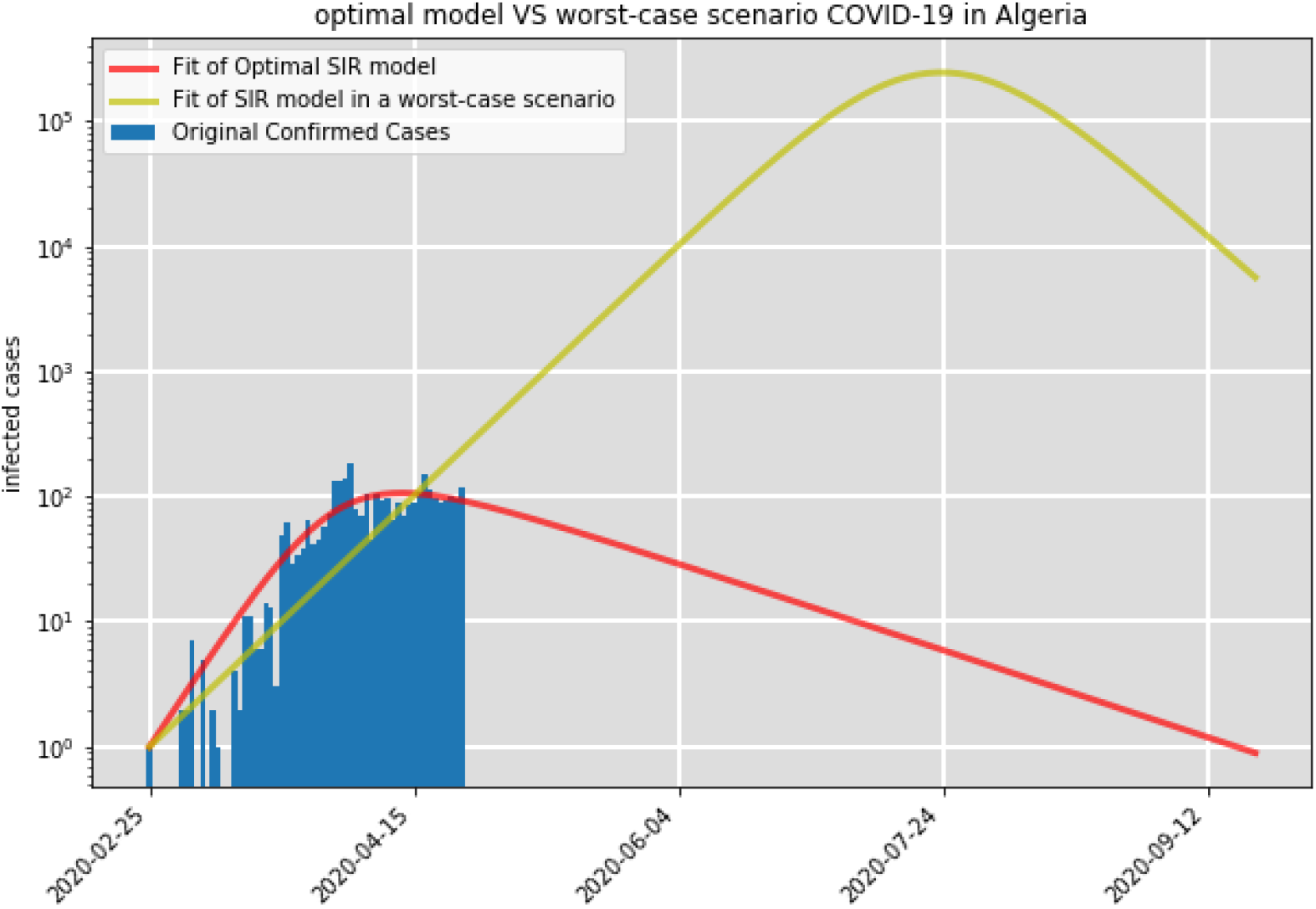
Prediction of the epidemic peak of COVID-19 in Algeria by fitting an optimal SIR model and an SIR model in a worst-case scenario.

## 4 Conclusions

In this study we predict the COVID-19 epidemic outbreak in Algeria using the number of daily infected cases, from 25 February 2020 to 24 April 2020. We have developed two SIR compartmental models without demographics, an optimal model and a model in a worst-case scenario. The estimated value of the basic reproduction number *R*_0_ using the optimal SIR model is 5.45, while *R*_0_ is estimated to be equal to 1.13 in the worst-case scenario COVID-19. *R*_0_ has a various determinants such as social behavior, population structure, viral evolution, environment, immunity as well as immigration policies, healthcare and Infrastructure of the health sector (2323, 22, 4). Comparing the prediction results of both models, we find that the COVID-19 epidemic peak in Algeria might attain 24 July 2020 with about 2444 ×10^2^ infections in a worst-case scenario. The end of this epidemic in Algeria is expected to be between 18 September 2020 and 16 November 2020, at the latest. Our findings agree with the results of Kuniya (17) in the case of COVID-19 outbreak in Japan, and with the WHO’s statement that COVID-19 will maybe disappear in the summer (17).

Following Kuniya (17), and based on the large differences in the predicted infections between the two models that represent the potentially preventable infections, we suggest that a strict containment strategy is needed in Algeria as soon as possible, over a relatively long period, in order to decrease the epidemic size.

In perspective, we will try to capture the effect of quarantine measures on COVID-19 outbreak in Algeria. For this, we would like to apply the SIQR model (Q stands for quarantined) (20).

## Data Availability

The data are obtained from:
https://github.com/CSSEGISandData/COVID-19/tree/master/csse_covid_19_data/csse_covid_19_time_series

